# The Impact of Gadopiclenol Dose on Myocardial and Left Ventricular Blood Pool T1 Shortening Times in Cardiac Magnetic Resonance Imaging

**DOI:** 10.1101/2025.06.18.25329760

**Authors:** Alexander Gashti, Joseph Bratsch, Michael Sanchez, Eric Linn, Richard Thompson

**Affiliations:** Health First Holmes Regional Medical Center, Melbourne, FL, United States

## Abstract

Gadopiclenol is a gadolinium-based contrast agent (GBCA) with high relaxivity, allowing for lower millimolar gadolinium (Gd) dosage while maintaining diagnostic image quality. While its efficacy in central nervous system (CNS) and body MRI has been studied, its application in cardiac MRI (CMR) remains largely unexplored. This raises important questions, especially given the known risks of Gd exposure, including nephrogenic systemic fibrosis (NSF) and Gd deposition disease (GDD). Due to the ability of all GBCAs to release trace amounts of free Gd, patients undergoing multiple contrast-enhanced MRI studies may face cumulative exposure risks. Identifying the lowest effective dose of gadopiclenol for CMR is essential, particularly for patients vulnerable to these conditions. T1 mapping offers a reproducible technique for evaluating myocardial tissue characteristics and was employed in this study to identify whether gadopiclenol 0.06 mmol/kg is an acceptable dose in CMR.

This observational, retrospective, pre-post contrast cohort study included 110 subjects (mean age: 65 years; 50 females, 60 males) with known or suspected cardiovascular lesions who underwent standardized CMR using Modified Look-Locker Inversion Recovery (MOLLI) T1 mapping on a 1.5 Tesla General Electric scanner. Native T1 maps were acquired in a midventricular short-axis slice and repeated 10 minutes post-contrast. A 0.06 mmol/kg dose of gadopiclenol was administered in 95% of patients. Myocardial and LV blood pool T1 values were measured pre- and post-contrast.

The mean myocardial T1 shortening was 608 ± 49 ms, reflecting a 60% reduction from baseline. The mean LV blood pool T1 shortening was 1244 ± 90 ms, corresponding to an 82% reduction. Images were technically adequate in 40% of patients, while 60% were technically difficult to achieve. The most common reason for technical difficulty was respiratory artifact, most often due to inconsistent breath holding. For most patients, technical difficulty did not impact T1 values. No infusion reactions were reported.

These findings support gadopiclenol’s potential as a lower-dose alternative to traditional GBCAs in CMR. Future research should examine its accuracy in extracellular volume (ECV) fraction estimation, performance at higher field strengths, and clinical utility in broader populations, including those with renal impairment. A cost-benefit analysis comparing gadopiclenol with conventional agents is also warranted.

## Background

CMR imaging relies on GBCAs to enhance visualization of myocardial tissue characteristics. Traditional GBCAs administered at standard millimolar Gd dosages achieve diagnostic image quality but raise safety concerns due to their association with NSF and GDD, particularly in patients undergoing multiple contrast-enhanced scans [1]. Gadopiclenol is a newer high-relaxivity GBCA designed to achieve imaging contrast at lower millimolar Gd dosage [2]. Although it has demonstrated strong imaging performance in CNS and body MRI and half the millimolar Gd dose [3], gadopiclenol’s role in CMR is largely unexplored. Reducing Gd burden without compromising diagnostic accuracy is especially relevant in CMR, where T1 mapping is used to quantitatively assess myocardial tissue relaxation dynamics and potentially detect a broad spectrum of myocardial pathologies.

This study aims to evaluate the impact of gadopiclenol at a mean dose of 0.06 mmol/kg on T1 relaxation times in the myocardium and LV blood pool at 1.5 Tesla. Our objective is to determine whether this lower dose achieves clinically meaningful T1 shortening while maintaining diagnostic imaging quality.

## Methods

This retrospective, observational cohort study included 110 patients (mean age: 65 years; 50 females, 60 males) with known or suspected cardiovascular lesions who were referred for CMR imaging within a 4-hospital health system. Given that all patients had previously underwent CMR imaging as part of standard clinical care, this retrospective study was exempt from institutional review board oversight. The primary outcome of the study was the T1 times in the myocardium and LV blood pool before and after contrast administration. The secondary outcome was the technical quality of the images as defined by the interpreting interventional cardiologist.

All patients underwent T1 mapping using a 1.5 Tesla MRI General Electric Healthcare scanner. Image acquisition and post-processing were performed using Circle Cardiovascular Imaging® software (cvi42) in conjunction with the institution’s Picture Archiving and Communication System (PACS) for image storage, retrieval, and management. These platforms were also used to document and track gadopiclenol administration in eligible patients. Patient data were retrospectively reviewed using the Sunrise Clinical Manager (SCM) electronic health record system.

Adult patients were included if they were eighteen years of age or older with known or suspected cardiovascular lesions if they could undergo the MOLLI breath holding sequence, and if they had no contraindications for CMR (metal implants, pacemakers, or severe claustrophobia). Patients were excluded if they had experienced allergic reaction to GBCAs in the past, if they were unable to cooperate with the breath holding sequence, or if they were pregnant or breastfeeding.

Baseline demographic and clinical data were collected, including age, gender, indication for the examination, type of study (stress or perfusion), gadopiclenol dose, power injection rate, and pre- and post-gadopiclenol myocardial and LV blood pool T1 times with corresponding standard deviations. T1 mapping was performed on a representative mid-ventricular short-axis slice for each patient using a MOLLI 3(3)3(3)5 sequence at 1.5 T. Native T1 images were acquired prior to contrast administration. Gadopiclenol was administered at a dose of 0.06 mmol/kg in 95% of patients. Post-contrast T1 maps were obtained following a 10-minute delay. T1 values were measured in both the myocardium and LV blood pool. Image quality was assessed by the interpreting interventional cardiologist and categorized as either (1) technically adequate or (2) technically difficult to achieve. For cases deemed technically difficult, the underlying reasons were further documented. Descriptive statistics were applied to the average T1 shortening time and the dispersion of T1 values around mean T1 times.

## Results

Baseline characteristics and CMR imaging data are summarized in Table 1. The mean gadopiclenol dose was 0.06 ± 0.004 mmol/kg, mean age 65 ± 15 years, mean BMI 29 ± 6, n=50 females, n=60 males. Average T1 values are reflected over the entire study population. Mean native myocardial T1 was 1010 ± 63 ms. Mean post contrast myocardial T1 402 ± 112 ms. Mean native LV blood pool T1 was 1506 ± 183 ms. Mean post contrast LV blood pool T1 was 263 ± 93 ms. T1 values for selected subgroups are further summarized on Table 2. Subgroup analysis reflecting T1 values are summarized for selected CMR indications within this population.

**Table 1.** Baseline Characteristics and CMR Imaging Data.

| **Baseline Characteristics and CMR Imaging Data Summary for All Pathologies** |  |
| --- | --- |
| Age, years | 65 ± 15 |
| Gender (male (n %)) | 60 (55%) |
| Body Mass Index kg/m<sup>2</sup> | 29 ± 6 |
| Imaging Modality | Stress 60 (55%)<br>Perfusion 50 (45%) |
| Gadopichlenol Dose (mmol/kg) | 0.06 ± 0.004 |
| Mean Native T<sub>1</sub> myocardium (msec) | 1010 ± 63 |
| Mean Native T<sub>1</sub> LV blood pool (msec) | 1506 ± 183 |
| Mean Post Contrast T<sub>1</sub> myocardium (msec) | 402 ± 112 |
| Mean Post Contrast T<sub>1</sub> LV blood pool (msec) | 263 ± 94 |

**Table 2.** Subgroup Analysis for Selected Groups.

| Row Labels | Patient Count | Mean Native T1 myocardium | Mean Post CA T1 myocardium | Mean Native T1 BP | Mean Post CA T1 BP |
| --- | --- | --- | --- | --- | --- |
| Amyloidosis | 3 | 1018 | 307 | 1545 | 244 |
| Arrhythmia | 16 | 991 | 389 | 1482 | 246 |
| Hypertrophic cardiomyopathy | 12 | 1011 | 369 | 1394 | 210 |
| Cardiomyopathy | 17 | 1006 | 455 | 1539 | 294 |
| Coronary artery disease | 17 | 1008 | 385 | 1507 | 285 |

Table 3 summarizes technical adequacy as defined by the primary reader. Imaging was deemed technically adequate in 45 patients, while the remainder faced technical difficulties. The most common reason for these difficulties was respiratory artifacts. Apart from combined motion and respiratory artifacts, technical difficulty did not impact T1 values. Two patients were excluded from the study lacking observable T1 values ascribed to a combination of respiratory and motion artifacts. The reason for the exam is summarized in Table 4. Scatterplots demonstrating the average pre-post contrast T1 values across the entire study population are shown on Figure 1. Typical T1 values are extrapolated from a single study on Figure 2.

**Table 3.**
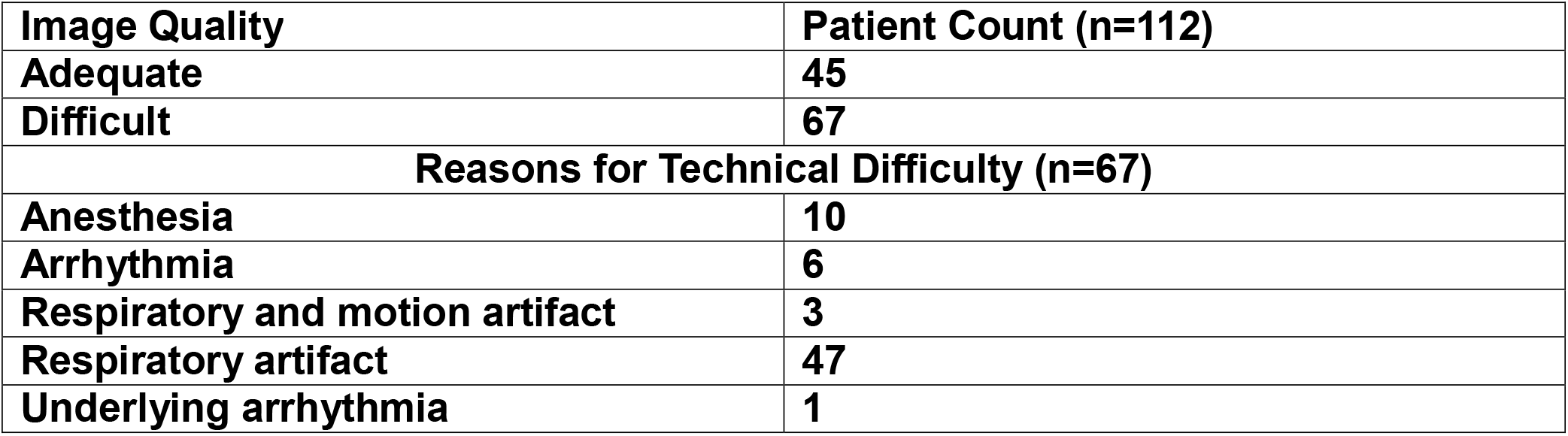
Technical Adequacy.

| Image Quality | Patient Count (n=112) |
| --- | --- |
| Adequate | 45 |
| Difficult | 67 |
| Reasons for Technical Difficulty (n=67) |  |
| Anesthesia | 10 |
| Arrhythmia | 6 |
| Respiratory and motion artifact | 3 |
| Respiratory artifact | 47 |
| Underlying arrhythmia | 1 |

**Table 4.**
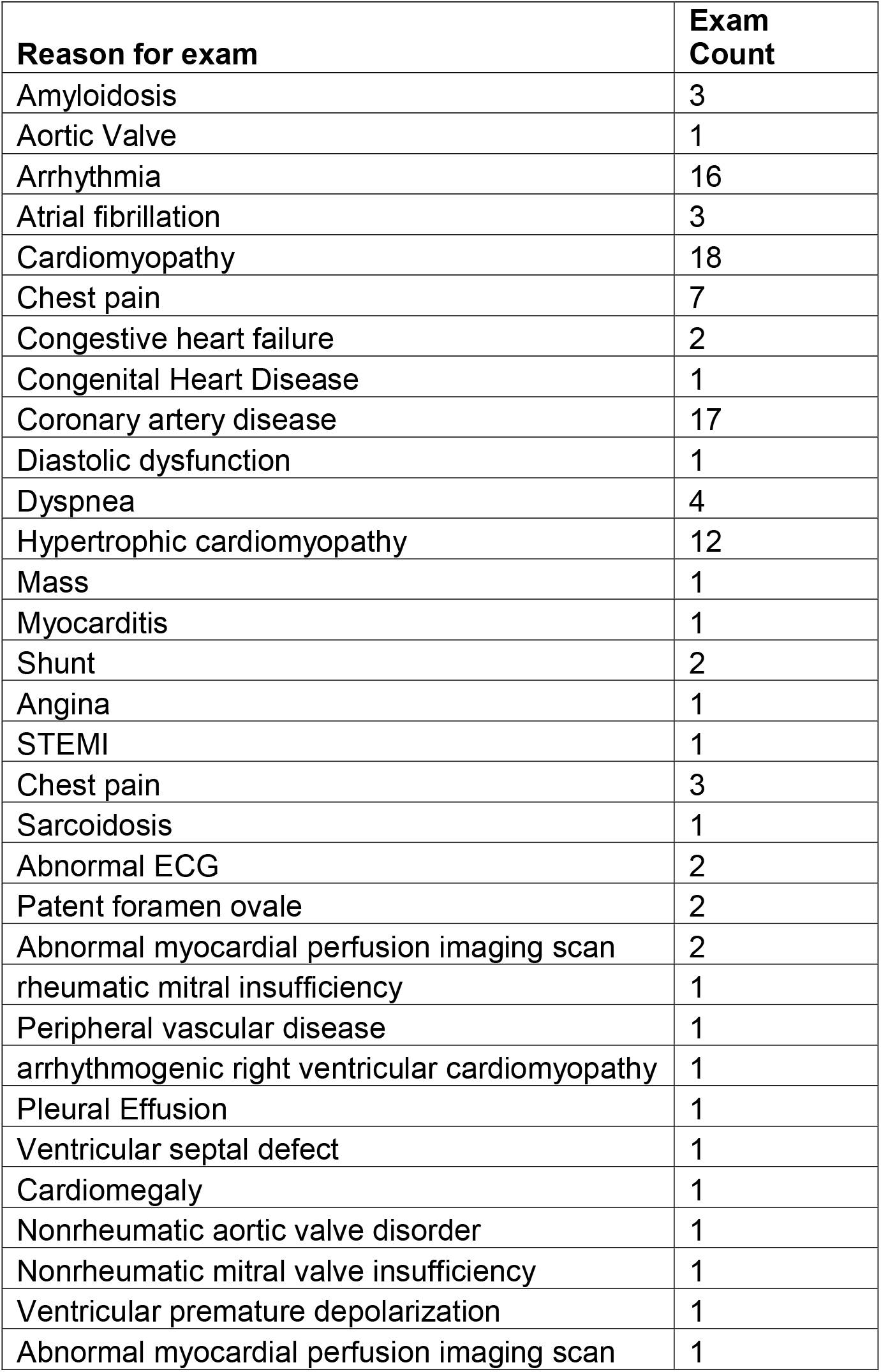
Reason for exam. The majority of CMR indications included cardiomyopathy (n=18), arrhythmia (n=19), hypertrophic cardiomyopathy (n=12), and coronary artery disease (n=17).

**Figure 1.**
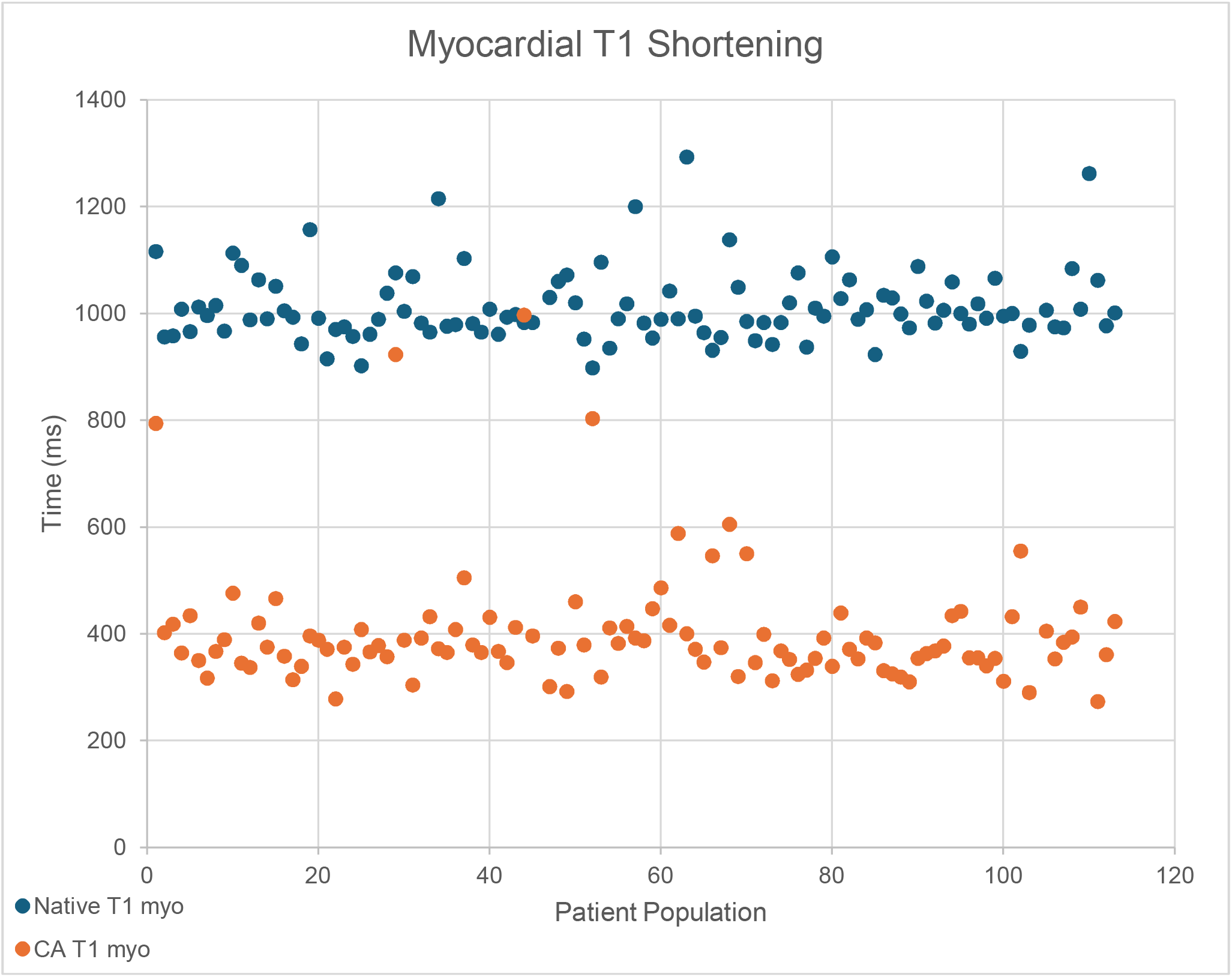
Average pre-post T1 Scatterplots

**Figure 1.**
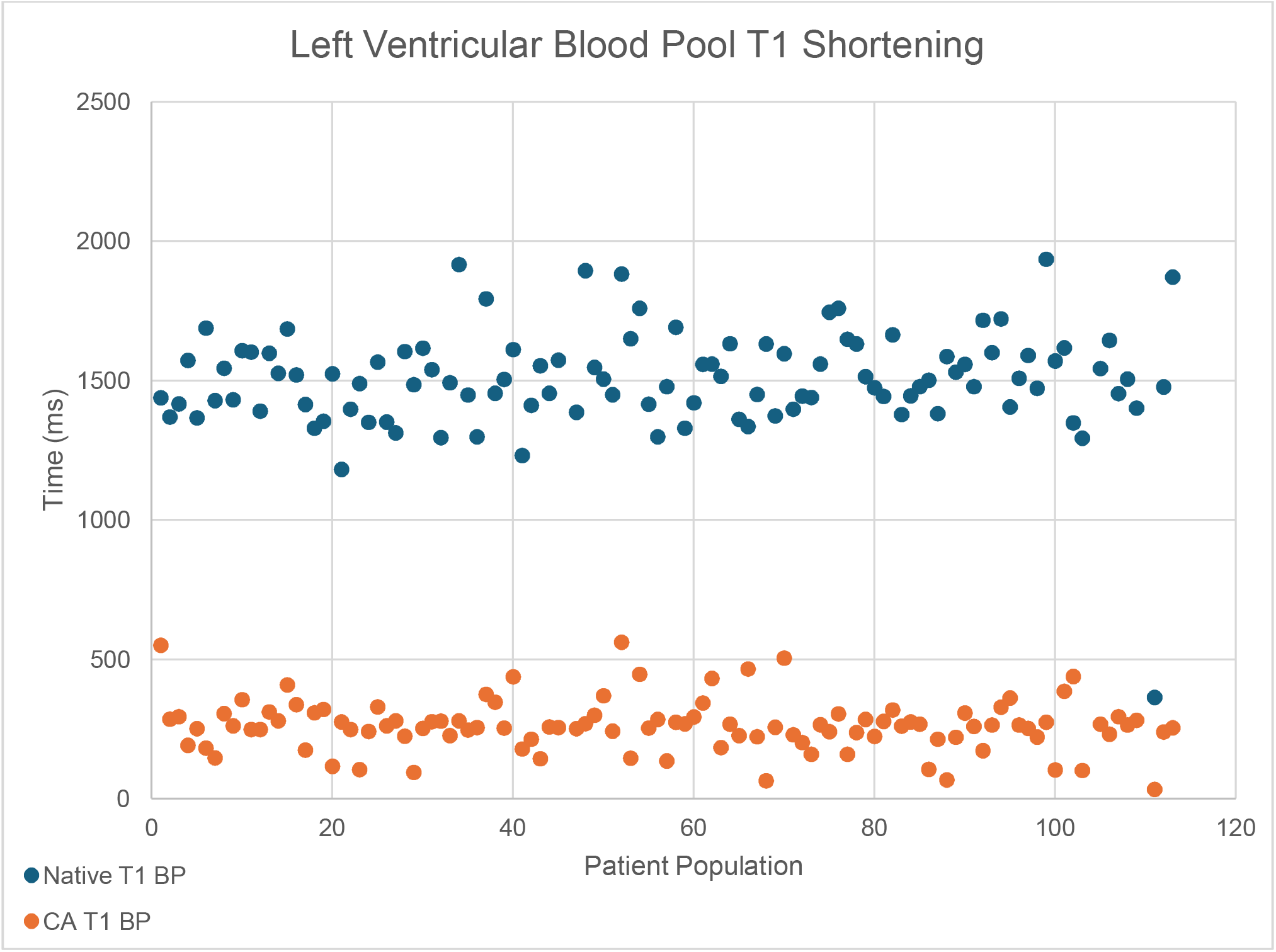
Gadopiclenol Enhanced Contrast Image: Acute Myocardial Infarction

**Figure 2.**
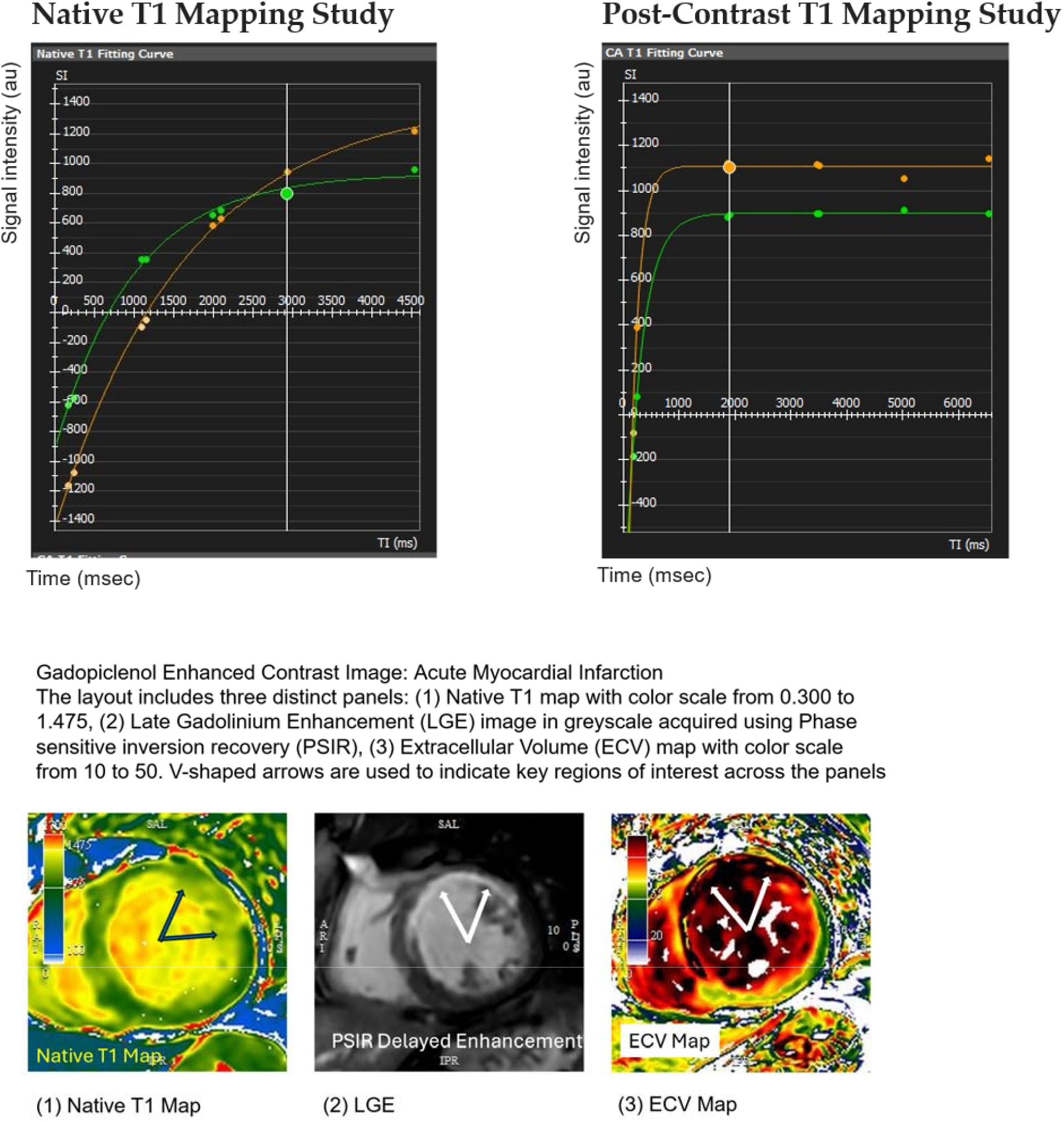
**Figure 2**. T1 shortening in a pre-to-post contrast T1 of the myocardium (in green) and LV blood pool (in orange) in a typical study of a single patient.

## Discussion

This study demonstrates gadopiclenol produces robust T1 shortening in both the myocardium and LV blood pool at routine post-contrast imaging intervals. The notably low millimolar Gd dose of 0.06 mmol/kg yielded substantial T1 shortening, suggesting dose reduction is feasible without compromising image quality. When comparing selected subgroups of this study to similarly designed studies, comparable T1 shortening is observed [4]. This study has several limitations. First, the absence of a comparator arm necessitated an indirect comparison between gadopiclenol and conventional GBCAs, which may limit the strength of the conclusions drawn. Second, image interpretation was performed by a single reader, potentially introducing bias and limiting generalizability. Additionally, certain MOLLI sequences that allow for shortened breath-hold durations (shMOLLI) were not included in the study protocol, specifically excluding patients imaged with shMOLLI sequences. Participants unable to sustain a breath-hold of at least 11 seconds were also excluded, which may introduce selection bias. Furthermore, the study was conducted exclusively at 1.5 Tesla, and the findings may not be generalizable to imaging performed at 3.0 Tesla. Finally, the retrospective and observational design of the study inherently limits the ability to establish causal relationships and may be subject to unmeasured confounding factors. Although T1 shortening was observed with gadopiclenol, the study did not evaluate its clinical implications, such as its effectiveness in lesion detection, fibrosis visualization, or diagnostic accuracy. Incorporating a qualitative assessment or limited quantitative comparison of lesion conspicuity relative to conventional GBCAs would provide valuable insight into its potential diagnostic utility.

When comparing pre- and post-contrast myocardial and left ventricular (LV) blood T1 values to those reported in similarly designed studies conducted at 1.5 Tesla—utilizing the same MOLLI sequence and collecting equivalent T1 indices—the values observed with gadopiclenol at a dose of 0.06 mmol/kg were found to be comparable to those obtained using higher millimolar doses of traditional GBCAs, including gadopentetate (0.15 mmol/kg) and gadobutrol (0.2 mmol/kg). This suggests that gadopiclenol may achieve similar T1 shortening effects at a lower dose, potentially offering a more efficient contrast-enhancement profile. These observations were initially made across a cohort with mixed cardiac pathologies. Following subgroup analyses, the T1 shortening effects observed with gadopiclenol closely mirrored those reported in comparably designed studies, given the use of significantly lower millimolar gadolinium doses— approximately half, or in some cases, one-third of the millimolar dose—compared to agents such as gadopentetate (0.15 mmol/kg) and gadobutrol (0.2 mmol/kg). This suggests that gadopiclenol may provide comparable T1 relaxation efficiency at reduced contrast agent concentrations, which could have implications for both safety and diagnostic performance.

The imaging protocol was well tolerated, with no reported infusion reactions among participants. MOLLI-based T1 mapping facilitated consistent image quality and reliable T1 quantification across the study cohort. These technical strengths likely contributed to the reproducibility of the results, with minimal influence from acquisition-related artifacts in the majority of cases. Diagnostic image quality was achieved in all participants, except in a few instances where significant respiratory and motion artifacts posed challenges. Despite these difficulties, image quality remained sufficient for diagnostic interpretation in all cases. Image quality was assessed by a single interventional cardiologist using a nominal scale (1 = adequate, 2 = technically difficult), with respiratory motion identified as the most frequent source of image degradation. To strengthen the objectivity and reproducibility of image quality assessment in future studies, the use of a semi-quantitative scale (e.g., a 5-point Likert-type scale) and the inclusion of inter-rater reliability measures—such as independent evaluation by an additional interventional cardiologist—are recommended.

While the sample size was modest, the study supports the potential for gadopiclenol to deliver diagnostic-quality T1 mapping at half the standard Gd dose. Further studies should investigate ECV quantification, myocardial scar detection, and other tissue characterization protocols.

**Table 1**. BMI – Imaging was attempted for a total of 110 patients. T1 values could not be obtained for 3 patients due to extreme technical difficulty during acquisition resulting from combined respiratory and motion artifacts.

## Data Availability

All data produced in the present study are available upon reasonable request to the authors

